# Experiences of accessing primary care by those living with long Covid in New Zealand

**DOI:** 10.1101/2025.04.29.25326651

**Authors:** Sarah Rhodes, Christina Douglas

## Abstract

**Background:** Long Covid is the persistence of symptoms beyond 12 weeks following acute Covid-19 infection. It is estimated to affect one in ten people and can be extremely debilitating. With few publicly funded long Covid clinics, most people rely on primary care providers as a first point of contact. There is currently limited understanding of the experience of accessing primary health care by adults living with long Covid in New Zealand.

**Purpose:** To explore the experiences of accessing primary health care by adults living with long Covid.

**Methods:** A narrative inquiry approach was used to capture participants’ lived experiences of accessing primary health care. Zoom interviews and discussions were conducted with study participants. The automatically generated transcripts were reviewed and corrected and the collated data were analysed using Braun and Clarke’s thematic analysis.

**Results:** Eighteen people participated in the interviews. Codes were identified and, through an iterative process, themes were generated, reviewed, and named. The seven themes included lack of upskilling of primary care staff; let down by the Government; self-advocacy and its cost; and throwing money at it.

**Conclusion(s):** The picture painted by participants was bleak with a sense that the world had moved on from Covid-19 and left them behind, with some experiencing a lack of support in primary health care. Better support might be achieved through a public awareness campaign for improved delivery of credible information, and greater utilisation of the allied health workforce.

## Introduction

Long Covid, or post-Covid condition, is the persistence of symptoms three months following an initial SARS-CoV-2 infection (1) and can be severely debilitating.(2) It is estimated to affect up to 45% of those who develop a Covid-19 infection (2) and the risk of long Covid persists with each subsequent Covid infection.(3) Although Covid-19 is no longer considered a public health emergency by the World Health Organization (4), long Covid presents an ongoing and complex challenge to affected individuals, their families and health systems.(5)

Although there is currently no cure for long Covid, there are ways of managing the condition. Studies show that various rehabilitation strategies (6) (7, 8) can result in a reduction in long Covid symptoms; as well as demonstrating some benefit from behavioural interventions.(9) Globally, both primary care providers and hospitals have developed services to provide long Covid management, with a range of options available, including in the United States (US) (10), the United Kingdom (UK) (11) and Australia.(12) However, research suggests that general practitioners (GPs) in some countries are impacted by the challenges of this new condition with its complex and varied clinical presentation (13), coupled with time-limited appointments.(14)

In New Zealand (NZ), the responsibility for managing long Covid sits in primary care, as highlighted by the Ministry of Health guidelines.(15) However, with a lack of funding and available resources to date, there is currently only one publicly available clinic, with a scattering of private providers across the country. With the primary care system under increasing pressure, and some general practices closed to enrolments due to underfunding, staff burnout and an ageing workforce, dedicated support for those living with long Covid looks unlikely.(16) Most people living with long Covid in NZ rely on their GP as a first point of contact.(13) However, there is a prevailing sense that NZ has moved on from the pandemic and that some of those living with long Covid have been overlooked, as is the case elsewhere.(17) This is particularly concerning for Māori and Pacific peoples, who already experience inequitable access to health care.(18) This concern about being left behind is echoed in the US (19) with concerns that minority communities are not well represented in long Covid research. Currently, little is known about patients’ experiences of accessing health care for long Covid or the quality of care received in NZ. The aim of our study was to explore the experiences of people living with long Covid in accessing primary care within NZ.

## Methods

This project was approved by the University of Otago Human Ethics Committee (H23/003). All procedures performed in studies were in accordance with the 1964 Helsinki declaration and its later amendments or comparable ethical standards. The study is reported in accordance with the Consolidated criteria for reporting qualitative research (COREQ) checklist.

### Participants

Eligible participants were anyone aged 18 or over with symptoms that met the World Health Organization definition of long COVID.(1)

Participants were recruited through convenience sampling via the NZ long haulers Facebook group, which has since become Long Covid Support Aotearoa (LCSA). Recruitment occurred between 1^st^ June 2023 and 1^st^ February 2024. This online community support group comprises over 2,600 members who are all people living with Long Covid; they are well informed and have been instrumental in raising awareness of long Covid within NZ. Written informed consent was obtained from each participant.

Drawing on the story telling tradition, the research used a narrative inquiry methodology, which sought to adopt a person-centred approach to give voice to each individual participant’s long Covid journey (20), as a means of understanding their lived experiences in depth. A strength of the narrative inquiry approach is that it provides flexibility in exploring layers of meaning both within individual stories and across multiple stories, allowing recognition of the uniqueness of each person’s reality, as well as the shared elements. This creates a richness to the data and explores the multiple dimensions of the journey, including the emotional aspects, associated with the impact of chronic illness. The aim was to provide a supportive and empowering space for these stories to be heard.(20) One potential limitation is that some participants can find the telling of their story both poignant and tiring.(21) This was anticipated and participants were aware that they could stop their story and/or take breaks from the interview/discussion group as they saw fit and without explanation.

The researchers each completed a reflexivity statement to identify their own reality and reflect on the individual values and beliefs they may hold. This allowed for better recognition of assumptions that they might bring to the research process, and to acknowledge how their subjectivity might shape the interview process and the subsequent analysis of transcripts (22), through their own knowledge and perspectives of long Covid. Participants were invited to take part in a group or individual Zoom session based on their preference. Participants were given the option to provide supplementary written information where the online sessions were considered too fatiguing. The aim was to recruit at least twelve people with lived experience. Each group session lasted approximately one hour.

The individual interviews and discussions explored participants’ experiences of living with long Covid. Part of this journey included experiences of accessing primary care for their condition. The focus was on providing space for peoples’ stories to be heard and deriving meaning from the collective experience. Zoom interviews began with an introduction to the study [Appendix 1] and a few loosely structured questions were used to encourage discussion about participants’ experiences [Appendix 2]. The zoom sessions were facilitated by a female member of the research team (SR) who holds a PhD and has previous experience of conducting qualitative interviews.

### Procedures

Participants’ demographic data were collected via email. Data were anonymized and collated before being stored on the primary researcher’s password protected computer.

### Data collection

Zoom interviews and discussions were recorded using the Zoom recording function, which generated a transcript that was temporarily stored in the Cloud. Once a transcript recording was completed and saved to the Cloud, the primary researcher received an automatically generated email. The transcript was then downloaded and saved on a password protected computer. The transcript was reviewed against the recording for accuracy, and the original recording was deleted. Transcripts were emailed to participants for any amendments, after which they were anonymised.

Where participants consented, but didn’t have the energy to participate in a Zoom session, they were given the opportunity to provide their answers to the questions in their own time and email them to the research team.

### Data analysis

The transcripts and email responses were analysed using Braun and Clarke’s six step thematic analysis.(23) This complimented the narrative inquiry approach to data collection, since it is an inductive approach which serves to identify and interpret patterns in the data, enabling development of a collective understanding of lived experiences. The researchers undertook familiarisation with the transcripts through repeated reading, making notes and developing preliminary codes. This was followed by a more robust coding process, identifying data into meaningful groups. Once codes were identified, they were reviewed and discussed, then analysed to generate larger over-arching themes [Appendix 3]. Themes were further refined through discussion and named to reflect their key quality. Write up involved reporting on the story the data reflected to present the findings in a meaningful way.

## Results and Discussion

Eighteen participants consented to be in the study. Sixteen were interviewed and two provided written responses. Participant characteristics can be seen in Table 1.

**Table 1:** Participant characteristics.

| Characteristics | Number (percentage) |
| --- | --- |
| **Gender** |  |
| Male | 3 (17) |
| Female | 13 (72) |
| Gender diverse | 2 (11) |
| **Ethnicity\*** |  |
| European | 15 (83) |
| Māori | 4 (22) |
| Pacific Peoples | 3 (17) |
| Asian | 0 (0) |
| Middle Eastern/Latin American/African | 1 (6) |
| Other Ethnicity | 0 (0) |
| *\*Some participants identified as more than one ethnicity* |  |
| **Age** |  |
| 18-29 | 3 (17) |
| 30-39 | 2 (11) |
| 40-49 | 4 (22) |
| 50-59 | 4 (22) |
| 60-69 | 4 (22) |
| 70+ | 1 (6) |
| **Urban/rural** |  |
| Urban | 10 (56) |
| Semi-rural | 2 (11) |
| Rural | 6 (33) |
| **Education** |  |
| Doctoral degree | 4 (22) |
| Master's degree | 7 (39) |
| Postgraduate diplomas and certificates, bachelor honours degree | 1 (6) |
| Bachelor's degree/graduate diplomas and certificates | 3 (17) |
| Diplomas | 2 (11) |
| Certificates | 1 (6) |

One participant responded with suggested amendments to their transcript.

Seven themes were developed: gaslighting and validation; lack of support and unmet need; inequity of available care; lack of upskilling of primary care staff; let down by the Government; self-advocacy and it’s cost; and throwing money at it.

### Theme 1: Gaslighting and validation

Our findings suggest that, for this cohort of people with lived experience of long Covid, there were several barriers to accessing care, including health professionals’ attitudes to the condition. Participants highlighted the fact that often their experiences of accessing primary care resulted in prejudice; primarily not being believed and being made to feel like they were wasting health professionals’ time, or being labelled as someone who didn’t contribute to society.

*One of the problems for people who’ve got long Covid that always makes me want to cry is that people don’t believe us*. [Participant 3]

*Someone who, effectively, my doctor’s term would be ‘a weight on society*.*’* [Participant 13]

This lack of validation of symptoms, and feeling dismissed by their doctors, has been an issue for many people presenting with long Covid in primary care, as reported by others living with the condition globally, including the UK and US. (14) (24, 25) (26)

This is despite greater understanding of the condition over time and a large body of literature confirming the physiological impacts.(27) This medical gaslighting may be partly due to the reliance on medical testing which does not always capture the physiological changes associated with long Covid, as well as an absence of established biomarkers to confirm the diagnosis.(27) Gaslighting of patients with ‘invisible’ symptoms is not new: it has been commonly experienced by the those living with conditions such as chronic pain for many years. (28)

Although most participants reported feeling stigmatized, some acknowledged that their experiences with health professionals were reassuring and helpful, reflecting the variation in attitudes within NZ. There was a sense that listening, acknowledging and being empathetic were valued, even in the absence of clear treatments.

*That validation is just so important like, I went to a new medical practitioner*, …, *last week. And the outstanding thing to me was, he said, ‘Oh, okay, how does long Covid affect you?’ And he talked about it as if it was perfectly normal to not be able to remember things*. [Participant 13]

Where health practitioners did listen and validate, this had a positive effect on the patient experience. Several studies have proposed that health professionals’ attitudes to long Covid present the greatest barrier to care.(29-31) Where there is continuity of care and a trusted relationship develops between patient and clinician, this can be game changing for the patient.(14, 31)

### Theme 2: Lack of support and unmet need

Participants raised the issue of facing barriers in the form of health professionals’ lack of supportive action. Several participants outlined the challenge of getting support in the form of long waiting times, no onward referral, and a lack of equipment or practical solutions, resulting in needs not being met.

*I can’t get any assistance. So, the doors keep closing everywhere*. [Participant 18]

*Go to the doctor and they say ‘Oh no, the waiting list is a year* [to see a specialist] [Participant 13]

Additionally, the lack of joined up care made wider support difficult to come by, with several participants underlining the constraints of the existing system.

*I struggled a lot with work and income around long Covid because they require all disabilities to have an end date and no one can give an end date when I’m going to stop having long Covid*. [Participant 4]

*Even people who are eligible for home help can’t get home help because of the way they fund the carers*. [Participant 1]

This experience of unmet need is not uncommon, with other studies highlighting delays in referral or treatment, as well as a lack of practical options offered.(32, 33) An online survey of 10,462 adults with long Covid, reported 34% found the health professional they accessed was unable to help them and 25% had a long waiting time to access help.(34) In other studies, those living with long Covid have highlighted available services as being slow, underdeveloped and not helpful. (35, 36) Overall, our study participants’ perceived lack of support left them feeling constrained by existing health system structures and that the system wasn’t fit for purpose. This is mirrored in other research where participants reported feeling that the system was set up for people to fail at obtaining help.(32)

### Theme 3: Inequity of available care

Health professionals’ lack of knowledge, lack of action and limited resources, contributed to a variation in care received across different locations and primary care providers, including evidence of urban-rural variation.

*It’s like so piecemeal, is the way I would describe it at the moment. You’re really lucky when you find someone good*. [Participant 8]

*I’m just putting my rural context on that because there’s nothing available here*. [Participant 1]

Sometimes the inequities were present even in the same city, which appeared to reflect a variation in primary care providers’ knowledge of both the condition and of the available referral pathways for long Covid related symptoms.

*[My GP’s] they haven’t known what the ***k to do with me. I know someone who’s had long Covid since she had Covid at the start of March this year; so only months. And she’s already got a POTS diagnosis. She’s already seen cardiology. She’s in the same city as me. She’s as sick as I am but she can get all of that stuff. I still can’t even get referred for a tilt table test*. [Participant 4]

One participant noted the variation in the care she had received compared with those who had more visible conditions, such a traumatic injuries.

*I almost wish I’d been in a car accident because then at least I would have some support*. [Participant 2]

Participants also commented on the fact that those with other visible health conditions, such as cardiovascular disease or type 2 diabetes, had better access to health services with established care pathways.(37) The invisible nature of long Covid appears to have contributed to the lack of available care.(27)

However, even differences between different invisible conditions were highlighted with some clinical presentations appearing to be more accepted and supported than others.

*If you don’t have a brain injury per se, but your brain is being affected by a virus, there just seems to be this silence around it*. [Participant 9]

The paucity of care received by those living with invisible illnesses, such as myalgic encephalomyelitis/chronic fatigue syndrome, has been well documented in the literature.(38, 39) However, some invisible conditions, such as concussion, do have well established treatment plans. Despite increasing recognition of the overlap between the clinical presentation of long Covid and persistent concussion, and calls in the literature to use post-concussion frameworks for those with long Covid (40), this appears to have gained limited traction in practice. Utilising existing skills to treat overlapping symptoms, as well as existing pathways as a starting point, may be one means of reducing inequity of care.

### Theme 4: Lack of upskilling of health care staff

The variation in services provided was perceived to be partly due to the lack of funding and resources to upskill health care staff in the assessment and management of long Covid. With fewer doctors entering general practice, the current primary care system in NZ is overwhelmed and unable to meet demand; existing GPs report feeling stressed and overworked,(41) leaving little time for upskilling. Participants wanted more explanation and guidance from the health professionals they saw regarding their condition.

*What I would love to be able to do is go to a medical professional and they could guide me through finding the right language to, to say what it is I am experiencing…* [Participant 14]

Other health professionals working in primary care were sometimes perceived as not having the skills to manage patients presenting with long Covid.

*She* [the HIP] *doesn’t know what to do. I feel so sorry for her. She’s I think, she’s overwhelmed*. [Participant 18]

This led to concerns that some health professionals were not accessing the current recommendations for management of long Covid, despite the publication of Long Covid rehabilitation guidelines by the Ministry of Health.(15)

*And there are still physios doing graded exercise therapy for people with long Covid. And, if you’re going to have physio referrals, they need, physios need updating and educating as well*. [Participant 15]

This sense that their health providers were still learning about the condition and were experiencing uncertainty regarding how to manage patients echoes the experiences of patients attending a post-Covid recovery clinic in Ohio.(42)

Experiences of staff having limited access to education and resources are also reflected in other study cohorts.(43) Although there is some evidence of resources being developed for healthcare workers to support patients with long Covid, (44) in reality, there does not appear to be a cohesive approach. Ultimately, dedicated workforce training on long Covid management, alongside clear signposting of credible online resources, is required to better support health professionals in primary care.(45)

### Theme 5: Let down by the Government

This inability to access the necessary care, and the perceived lack of funding and support for staff in primary care to upskill, led to a feeling of being let down by the Government.

*Everyone goes ‘oh the poor [GP] practice, you know, the poor practice, they are so under pressure, it’s like, well stuff it actually! You know, this is where you’re meant to be coping with us. The Government has said you are responsible for this chronic illness but they haven’t got the resources to do it*. [Participant 8]

Additionally, there was disappointment and anger at what was regarded as a lack Government action.

*So it’s greatly ironic that I’m actually here today and extremely angry because everything I planned for* [in terms of policy] *has been ignored by the public policy system*. [Participant 10]

*I mean, I feel totally pissed off with the Government,…, But for the Government to say, to be so silent on long Covid. I mean, it’s not just, it’s illegal*. [Participant 11]

Long Covid is associated with increased health care use and substantial primary care costs.(46) There is a perceived lack of government action in NZ, in terms policy initiatives and additional funding to primary care, to support those living with long Covid. With the NZ health system under increasing pressure and an admission that the system is in crisis with high levels of unmet need (47), specific funding for long Covid is unlikely. The current situation is one of geographical variation and fragmented care, primarily offered by private providers. This mirrors experiences elsewhere. In Ontario, Canada, the lack of a cohesive long Covid strategy by government has led to a disjointed approach to patient care, which is now under threat.(48) However, this is not the case everywhere. In England, the NHS has invested £34 million on over 80 adult long Covid clinics.(11) Likewise, most top hospitals in the US provide some form of long Covid service, although there is variation in terms of what is offered.(10)

In other places, the absence of clear government action has resulted in strong advocacy from the primary care sector, with organisations such as the Royal Australasian College of Physicians raising concerns about the closure of Australian long Covid clinics and appealing for government funding.(49) Studies have highlighted an urgent need for innovative, cost effective models of care to successfully meet the needs of patients living with long Covid.(12)

### Theme 6: Self advocacy and its cost

In the absence of readily available support, self-advocacy was a key feature of most participants’ approach to accessing care. Some viewed this as a necessary part of the process to move things forward in getting the support they required. They were proactive and not afraid to ask for what they needed.

*My doctor wouldn’t refer me for ages. So I basically got the clinic to contact her. I’d given them permission to look at my medical records. So I went over her head to do it*. [Participant 12]

However, others expressed their frustration and annoyance that it had to be this way.

*You know, like all that work, all that advocacy that I have to do, all of that in order to get them to listen is infuriating*. [Participant 9]

For others, self-advocacy represented them finding their own solutions to managing their condition or sending information to health professionals.

*I think we’ve all had to do a lot of investigation into stuff because when there is no one else that’s had it and, you know, with my GP I was constantly sending her links; please read this, you know*. [Participant 1]

This need to find their own solutions came at a cost.

*I’ve got to read and learn myself about how I could then apply it to my schedule. But there is a huge energy cost to that*. [Participant 12]

Patients seeking their own creative solutions to managing their condition is not unique to NZ.(30) Elsewhere, others have been proactive in their engagement with the health system (36) through decisive action such as switching GPs, demanding referral to a long Covid clinic or asking for referral to specialist.(14) A 2023 UK study reported that patients viewed advocating for themselves as an attempt to regain control in a situation of uncertainty.(26)

This proactive approach to health care by patients has the potential to shape patient-clinician partnerships in the future. The notion of shared decision making is at the heart of person-centred care and acknowledges the patient’s expertise about their condition.(50) However, nurturing therapeutic relationships is challenging in a system where GPs are overstretched due to staff shortages, an ageing workforce and an increasing number of patients presenting with complex conditions.(51) One solution might be to better utilise the allied health workforce in the primary care sector, which aligns with the existing NZ Government priorities.(52)

### Theme 7: Throwing money at it

The lack of recognised treatments for long Covid led to people seeking out alternative options; some without any supporting evidence and often at a high financial cost.

*I’ve tried over 20 different forms of treatment and therapy*. [Participant 7]

*So she told me the name of the expert, …, so I made a private appointment to see him at $405. Just about killed me. But anyway, and he had nothing to offer. He had nothing to offer*. [Participant 3]

Participants acknowledged that they were prepared to try any form of alternative treatment in an attempt to alleviate symptoms.

*I’ve tried the alternative, you know. Hypobaric chambers, osteopaths, various different supplements, acupuncture, just in desperation to do something*. [Participant 2]

*You can spend an awful lot of money on alternative treatments…There’s lots of things you could spend your money on and you’d still be going, I’m not sure if that’s helping*. [Participant 8]

There was frustration that people with long Covid were ripe for exploitation due to the failure of health services to provide for them.

*Online it is clear that desperate souls are open to being taken advantage of by alternative practitioners – our care should be clear in mainstream medicine so we are not open to this!* [Participant 18]

Although there is little available research to support this happening in other long Covid cohorts, one study highlighted the impact of people with long Covid engaging with dubious health sources online.(14) The issue of “quackery” in health care is not new, with a recent scoping review seeking to determine the reason for the development of quack medicine.(53) The authors’ concluded that quack medicine is caused by a range of factors, including political, economic, sociocultural and psychological.(53) In the context of long Covid, it is apparent that this population are vulnerable due to the ongoing gaslighting some have experienced at the hands of medical professionals (14) and the lack of available treatments.(27) The resulting disappointment and despair (53) provides a perfect opportunity for exploitation by unscrupulous practitioners.

Insufficient information and health literacy both contribute to susceptibility to quackery.(53) Raising awareness of long Covid through a public awareness campaign, with a particular focus on those groups who are often marginalised by health care, might serve as a good starting point. This could include communicating where to access credible online resources, which could support some people in self-managing their condition. The need for better access to information has been highlighted by those living with long Covid in other studies.(54)

### Strengths and limitations

One of the key strengths of this study was the member checking of transcripts to ensure they accurately reflected the patient voice. A key limitation of our study was that participants were all recruited from an online support group and their views may not reflect the wider cohort of people living with long Covid. Another limitation was the lack of diversity within our sample in terms of educational background; most participants were highly educated and had a post graduate qualification. Therefore, our findings may not be representative of the wider population affected by long Covid and cannot be generalised.

## Conclusion

This is the first study to highlight the experiences of accessing primary care by a cohort of people living with long Covid in NZ. The picture painted by these participants is bleak with a sense that the world had moved on from Covid-19 and left them behind. Despite the existence of long Covid for several years, and the Ministry of Health providing clinical recommendations, there is still only one publicly funded long Covid clinic in NZ. Patients dissatisfied with the current system experience challenges, including gaslighting, unmet need, inequity of care and uncertainty amongst health providers regarding their condition. In response they are having to self-advocate strongly which often comes at both a high personal and financial cost. With the current pressure on the NZ health system, any funding to provide further public services for long Covid looks increasingly unlikely. Potential cost-effective solutions to better support those living with the condition include a public awareness campaign for better delivery of credible information to support self-management where possible, and better utilisation of the existing allied health workforce, to lessen the pressure on primary care providers.

## Data Availability

All relevant data are within the manuscript and its Supporting Information files.

## Acknowledgements

We wish to acknowledge the School of Physiotherapy, University of Otago for providing access to resources and support. We also wish to acknowledge the participants who entrusted their stories to us and shared their journeys.

## References

1. World Health Organization. Post covid-19 condition (long covid) 2022. Available from: https://www.who.int/europe/news-room/fact-sheets/item/post-covid-19-condition#:~:text=It%20is%20defined%20as%20the,months%20with%20no%20other%20explanation.

2. O’Mahoney LL, Routen A, Gillies C, Ekezie W, Welford A, Zhang A, et al. The prevalence and long-term health effects of Long Covid among hospitalised and non-hospitalised populations: a systematic review and meta-analysis. eClinicalMedicine. 2023;55.

3. Bosworth ML, Shenhuy B, Walker AS, Nafilyan V, Alwan NA, O’Hara ME, et al. Risk of New-Onset Long COVID Following Reinfection With Severe Acute Respiratory Syndrome Coronavirus 2: A Community-Based Cohort Study. Open Forum Infect Dis. 2023;10(11):ofad493.

4. Sarker R, Roknuzzaman ASM, Hossain MJ, Bhuiyan MA, Islam MR. The WHO declares COVID-19 is no longer a public health emergency of international concern: benefits, challenges, and necessary precautions to come back to normal life. Int J Surg [Internet]. 2023 Sep 1 PMC10498846]; 109(9):[2851-2 pp.]. Available from: https://www.ncbi.nlm.nih.gov/pubmed/37222700.

5. Parotto M, Gyöngyösi M, Howe K, Myatra SN, Ranzani O, Shankar-Hari M, et al. Post-acute sequelae of COVID-19: understanding and addressing the burden of multisystem manifestations. The Lancet Respiratory Medicine. 2023;11(8):739–54.

6. Zheng C, Chen XK, Sit CH, Liang X, Li MH, Ma AC, et al. Effect of Physical Exercise-Based Rehabilitation on Long COVID: A Systematic Review and Meta-analysis. Med Sci Sports Exerc. 2024;56(1):143–54.

7. Nopp S, Moik F, Klok FA, Gattinger D, Petrovic M, Vonbank K, et al. Outpatient Pulmonary Rehabilitation in Patients with Long COVID Improves Exercise Capacity, Functional Status, Dyspnea, Fatigue, and Quality of Life. Respiration. 2022;101(6):593–601.

8. Pouliopoulou DV, Macdermid JC, Saunders E, Peters S, Brunton L, Miller E, et al. Rehabilitation Interventions for Physical Capacity and Quality of Life in Adults With Post-COVID-19 Condition: A Systematic Review and Meta-Analysis. JAMA Netw Open. 2023;6(9):e2333838.

9. Zeraatkar D, Ling M, Kirsh S, Jassal T, Shahab M, Movahed H, et al. Interventions for the management of long covid (post-covid condition): living systematic review. Bmj. 2024;387:e081318.

10. Haslam A, Prasad V. Long COVID clinics and services offered by top US hospitals: an empirical analysis of clinical options as of May 2023. BMC Health Services Research. 2024;24(1):684.

11. Greenhalgh T, Darbyshire J, Ladds E, Van Dael J, Rayner C. Working knowledge, uncertainty and ontological politics: An ethnography of UK long covid clinics. Sociology of Health & Illness. 2024;46(8):1881–900.

12. Luo S, Zheng Z, Bird SR, Plebanski M, Figueiredo B, Jessup R, et al. An Overview of Long COVID Support Services in Australia and International Clinical Guidelines, With a Proposed Care Model in a Global Context. Public Health Rev. 2023;44:1606084.

13. Brode WM, Melamed E. A practical framework for Long COVID treatment in primary care. Life Sciences. 2024;354:122977.

14. Turk F, Sweetman J, Chew-Graham CA, Gabbay M, Shepherd J, van der Feltz-Cornelis C. Accessing care for Long Covid from the perspectives of patients and healthcare practitioners: A qualitative study. Health Expect. 2024;27(2):e14008.

15. Health Mo. Clinical Rehabilitation Guideline for People with Long COVID (Coronavirus Disease) in Aotearoa New Zealand: Revised December 2022. Wellington 2022 13 December 2022.

16. Andrew A. Aotearoa New Zealand general practice workforce crisis: what are our solutions? J Prim Health Care. 2024;16(2):214–7.

17. Evered JA, LaJeunesse A, Wynn M, Mrig E, Schlesinger M, Grob R. Gaps in benefits, awareness, and comprehension that leave those with long COVID vulnerable. Chronic Illness. 2023:17423953231210117.

18. Harris RB, Cormack DM, Stanley J. Experience of racism and associations with unmet need and healthcare satisfaction: the 2011/12 adult New Zealand Health Survey. Aust N Z J Public Health. 2019;43(1):75–80.

19. Medeiros M, Edwards HA, Baquet CR. Research in the USA on COVID-19’s long-term effects: measures needed to ensure black, indigenous and Latinx communities are not left behind. J Med Ethics. 2023;49(2):87–91.

20. Weiss CR, Johnson-Koenke R. Narrative Inquiry as a Caring and Relational Research Approach: Adopting an Evolving Paradigm. Qualitative Health Research. 2023;33(5):388–99.

21. Wang CC, Geale SK. The power of story: Narrative inquiry as a methodology in nursing research. International Journal of Nursing Sciences. 2015;2(2):195–8.

22. Olmos-Vega FM, Stalmeijer RE, Varpio L, Kahlke R. A practical guide to reflexivity in qualitative research: AMEE Guide No. 149. Med Teach. 2022:1–11.

23. Braun V, Clarke V. Using thematic analysis in psychology. Qualitative Research in Psychology [Internet]. 2006; 3:[77–101 pp.]. Available from: 10.1191/1478088706qp063oa.

24. Au L, Capotescu C, Eyal G, Finestone G. Long covid and medical gaslighting: Dismissal, delayed diagnosis, and deferred treatment. SSM Qual Res Health [Internet]. 2022 Dec PMC9448633]; 2:[100167 p.]. Available from: https://www.ncbi.nlm.nih.gov/pubmed/36092770.

25. Owen R, Ashton RE, Skipper L, Phillips BE, Yates J, Thomas C, et al. Long COVID quality of life and healthcare experiences in the UK: a mixed method online survey. Qual Life Res. 2024;33(1):133–43.

26. Skilbeck L, Spanton C, Paton M. Patients’ lived experience and reflections on long COVID: an interpretive phenomenological analysis within an integrated adult primary care psychology NHS service. J Patient Rep Outcomes. 2023;7(1):30.

27. Davis HE, McCorkell L, Vogel JM, Topol EJ. Long COVID: major findings, mechanisms and recommendations. Nat Rev Microbiol [Internet]. 2023 Mar PMC9839201]; 21(3):[133–46 pp.]. Available from: https://www.ncbi.nlm.nih.gov/pubmed/36639608.

28. Quintner J. Why Are Women with Fibromyalgia so Stigmatized? Pain Med. 2020;21(5):882–8.

29. Ladds E, Rushforth A, Wieringa S, Taylor S, Rayner C, Husain L, et al. Persistent symptoms after Covid-19: qualitative study of 114 “long Covid” patients and draft quality principles for services. BMC Health Services Research. 2020;20(1):1144.

30. Ladds E, Rushforth A, Wieringa S, Taylor S, Rayner C, Husain L, et al. Developing services for long COVID: lessons from a study of wounded healers. Clinical medicine. 2021;21(1):59–65.

31. Leggat FJ, Heaton-Shrestha C, Fish J, Siriwardena AN, Domeney A, Rowe C, et al. An exploration of the experiences and self-generated strategies used when navigating everyday life with Long Covid. BMC Public Health. 2024;24(1):789.

32. McNabb KC, Bergman AJ, Smith-Wright R, Seltzer J, Slone SE, Tomiwa T, et al. “It was almost like it’s set up for people to fail” A qualitative analysis of experiences and unmet supportive needs of people with Long COVID. BMC Public Health. 2023;23(1):2131.

33. Baz SA, Fang C, Carpentieri JD, Sheard L. ‘I don’t know what to do or where to go’. Experiences of accessing healthcare support from the perspectives of people living with Long Covid and healthcare professionals: A qualitative study in Bradford, UK. Health Expectations. 2023;26(1):542–54.

34. Brus IM, Spronk I, Polinder S, Loohuis A, Tieleman P, Heemskerk SCM, et al. Self-perceived barriers to healthcare access for patients with post COVID-19 condition. BMC Health Serv Res. 2024;24(1):1035.

35. Hossain MM, Das J, Rahman F, Nesa F, Hossain P, Islam AMK, et al. Living with “long COVID”: A systematic review and meta-synthesis of qualitative evidence. PLoS One. 2023;18(2):e0281884.

36. Macpherson K, Cooper K, Harbour J, Mahal D, Miller C, Nairn M. Experiences of living with long COVID and of accessing healthcare services: a qualitative systematic review. BMJ Open. 2022;12(1):e050979.

37. Bellary S, Kyrou I, Brown JE, Bailey CJ. Type 2 diabetes mellitus in older adults: clinical considerations and management. Nature Reviews Endocrinology. 2021;17(9):534–48.

38. Sowińska A, Pezoa Tudela R. Living with invisible medical disabilities: experiences and challenges of Chilean university students disclosed in medical consultations. Int J Qual Stud Health Well-being. 2023;18(1):2221905.

39. Cadle B, Scholtz M. Notions of the visible and the invisible: visualising the otherness of women with invisible (gynaecological) illness. Visual Studies.1–11.

40. Davidson BS, Noteboom L, Pierro H, Kantor C, Stoot D, Stoot F, et al. Post-Concussion Assessment as a diagnostic and mechanistic framework for treating patients with Long COVID. medRxiv. 2022:2022.09.24.22280310.

41. Betty B, Scott-Jones J, Toop L. State of general practice in New Zealand. The New Zealand Medical Journal (Online). 2023;136(1582):8–10.

42. MacEwan SR, Rahurkar S, Tarver WL, Forward C, Eramo JL, Teuschler L, et al. Patient Experiences Navigating Care Coordination For Long COVID: A Qualitative Study. J Gen Intern Med. 2024;39(8):1294–300.

43. Pinto Pereira SM, Newlands F, Anders J, Banerjee A, Beale S, Blandford A, et al. Long COVID: what do we know now and what are the challenges ahead? Journal of the Royal Society of Medicine. 2024;117(7):224–8.

44. Leighton J, Reis L, Wasilewski M, Simpson R. Showcasing a ‘Long-COVID (LC) Workforce Toolkit’ for Health and Social Care Providers: Providing resources, supports, and a mapping of networks that support rehabilitative efforts. Archives of Physical Medicine and Rehabilitation. 2024;105(4):e10.

45. Hodgson CL, Broadley T. Long COVID—unravelling a complex condition. The Lancet Respiratory Medicine. 2023;11(8):667–8.

46. Tufts J, Guan N, Zemedikun DT, Subramanian A, Gokhale K, Myles P, et al. The cost of primary care consultations associated with long COVID in non-hospitalised adults: a retrospective cohort study using UK primary care data. BMC Prim Care. 2023;24(1):245.

47. Keene L, Wild H, Mills V. Anatomy of a health crisis. The New Zealand Medical Journal (Online). 2024;137(1594):9–12.

48. A CLJ. Lack of Ontario long COVID strategy risks care: ministry documents The Canadian Press. 2023.

49. Jolyon A. Call for more investment in long COVID treatment. News GP. 2023.

50. Practitioners TRCoG. The power of relationships: what is relationship-based care and why is it important? General practice COVID-19 recovery. London Royal College of General Practitioners 2021 June 2021.

51. N GRBJB. Quantifying and understanding the impact of unmet need on New Zealand general practice. Dunedin: University of Otago; 2024 4 June 2024.

52. Hauora MoHM. Hauora Haumi Allied Health Report 2024. Wellington: Minsitry of Health; 2024 27 June 2024.

53. Amir-Azodi A, Setayesh M, Bazyar M, Ansari M, Yazdi-Feyzabadi V. Causes and consequences of quack medicine in health care: a scoping review of global experience. BMC Health Serv Res. 2024;24(1):64.

54. Razai MS, Al-Bedaery R, Anand L, Fitch K, Okechukwu H, Saraki TM, et al. Patients’ Experiences of “Long COVID” in the Community and Recommendations for Improving Services: A Quality Improvement Survey. Journal of Primary Care & Community Health. 2021;12:21501327211041846.

